# Associations between commonly consumed beverages and the oral microbiome in three US cohort studies

**DOI:** 10.64898/2026.10.07.26364990

**Authors:** Lynn E. Ferro, Samuel Anyaso-Samuel, Yukiko Yano, Xing Hua, Yunhu Wan, Vicky C. Chang, Casey L. Dagnall, Kristine Jones, Belynda D. Hicks, Amy Hutchinson, Linda M. Liao, Wen-Yi Huang, Neal D. Freedman, Laura E. Beane Freeman, Dale P. Sandler, Christian C. Abnet, Jianxin Shi, Rashmi Sinha, Erikka Loftfield, Emily Vogtmann

## Abstract

Multiple external factors shape human microbial communities. Beverages represent particularly frequent dietary exposures that may impact oral microbiome composition. We examined associations of coffee, tea, alcohol, sugar-sweetened beverages, artificially-sweetened beverages, and fruit juice consumption with the oral microbiome in three large cohort studies (n=5,598). Beverage consumption was assessed by food frequency questionnaire and categorized as non-consumers and tertiles of intake among consumers, except fruit juice, which was categorized into quartiles. DNA from oral wash specimens was sequenced (16S rRNA, V4 region). Associations with alpha and beta diversity and individual genera were assessed using multivariable linear, logistic, count-based regression models and weighted permutation-based models accounting for multiple comparisons in genus-level analyses. Only alcoholic beverage consumption was associated with all alpha diversity metrics; the highest tertile of alcohol drinking was associated with 10.18 (95% CI: 5.56, 14.80) greater observed amplicon sequence variants vs. non-consumers. All beverages accounted for 4.06% of variability in beta diversity (unweighted UniFrac); in univariable models, alcoholic beverages had the largest R² among individual beverages (0.86%) and accounted for the majority (25/28, 89.3%) of genus-level associations, mostly with beer and liquor. For example, high alcoholic beverage intake (T3 vs. non-consumers) was associated with 2.25 and 2.17 times the odds of having *Megasphaera* and *Peptostreptococcus*, respectively, and 23% lower abundance of *Lautropia*. Alcoholic beverage consumption was strongly associated with oral microbes, including taxa linked to poor oral health; these findings suggest that alcohol-associated differences in the oral microbiome may have implications for oral and systemic health.

## Introduction

The oral microbiome is a community of microorganisms, including bacteria, fungi, viruses, and archaea, that reside in the oral cavity.^1,2^ Oral bacteria have metabolic and immune-related functions such as protection of the oral cavity, digestion, immune response, and prevention of disease.^3,4^ The oral microbiome plays a large role in oral health, specifically due to the microbial mechanisms by which dental caries and periodontal disease develop, in addition to its contribution to overall host health.^1^ Further, the oral microbiome has been linked to systemic diseases including Alzheimer’s disease, diabetes, cardiovascular disease, inflammatory bowel disease, obesity, and some cancers,^1^ particularly lung, colorectal, pancreatic, and head and neck cancers.^5–8^

The composition of the oral microbiome has been observed to be impacted by maternal, environmental, and lifestyle factors including dental hygiene practices, medication use, and diet.^9,10^ Beverages may be an integral dietary contributor due to their frequent, habitual and longer duration of exposure compared to foods which are consumed more episodically. According to NHANES 2017-2018 data, adults consume approximately 84 fluid ounces of beverages, about 10.5 cups, per day where approximately half is water.^11^ However, roughly half of the calories one consumes from beverages alone were from alcoholic and sweetened beverages,^11^ and these beverages are largely associated with increased mortality and cardiovascular risks.^12^ Moreover, the impact of beverages, specifically those higher in sugar may be profound due to alterations in the oral microbiome and frequent, widespread exposure throughout the oral cavity.^13–15^ Coffee, tea, alcoholic beverages, sugar sweetened beverages (SSBs), artificially sweetened beverages (ASBs), and fruit juice are all commonly consumed beverages in the United States.^11^ A prior cross-sectional analysis of the Cancer Prevention Study II (CPS-II) and the Prostate, Lung, Colorectal, and Ovarian (PLCO) Cancer Screening Trial found no association between coffee consumption and the oral microbiome, but found increased diversity and differences in bacterial community composition for tea drinkers.^16^ Studies examining alcohol have found that higher consumption is associated with increased alpha diversity and differences in bacterial composition.^17–19^ While evidence suggests a positive relationship of SSB consumption with dental caries and erosion,^20^ little research has explored the relationship of SSBs, ASBs, and fruit juice with the oral microbiome. However, in our previous investigation of the Healthy Eating Index (HEI) and the oral microbiome, while no association was found with diet quality overall, we observed an association with the added sugar component of the HEI and the oral microbiome.^21^ Given that SSBs may be a major source of added sugar in one’s diet and contributes an average of 24% of daily added sugar intake,^22^ it is possible that this previously observed association was at least partially driven by SSB consumption.

We evaluated the influence of beverage consumption on the diversity and composition of the oral microbiome, we examined the associations of coffee, tea, alcoholic beverages, SSBs, ASBs, and fruit juice with the oral microbiome using data from oral microbiome studies nested within the Agricultural Health Study (AHS), the NIH-AARP Diet and Health Study, and PLCO.

## Methods

### Study population

We previously conducted a nested case-cohort study of the oral microbiome and cancer risk within three US prospective cohort studies: AHS (NCT00352924), NIH-AARP (NCT04750915), and PLCO (NCT00339495). Details of the study design and methods of the nested case-cohort study have been described elsewhere.^7,8^ Briefly, a subset of participants from AHS, NIH-AARP, and the PLCO control arm provided oral wash specimens in 1999-2003, 2005, and 2000-2003, respectively. Participants with oral wash specimens who developed incident lung, colorectal, esophageal, head/neck, stomach, small intestine, pancreatic, or hepatobiliary tract cancer were identified along with a comparison referent sub cohort based on age, sex, and smoking status categories. Due to the sampling design, incident cancer cases were assigned a sampling weight of 1 and weights for those in the referent sub-cohort were based on the inverse of the observed sampling fractions.

We excluded participants with missing dietary data (n=604), microbiome data (n=11), a prevalent cancer at the time of oral wash collection (n=424), Parkinson’s disease (n=422), or implausible caloric intake (<600 kcal or > 5,000 kcal) (n=148). After exclusions, we included 1,468 participants from the AHS cohort (weighted N = 26,752), 1,934 participants from the NIH-AARP cohort (weighted N = 25,084), and 2,196 participants from the PLCO cohort (weighted N = 34,957). All studies received approval from their respective institutional review boards, and all participants provided written informed or implied consent.

### Dietary assessment

A validated food frequency questionnaire (FFQ) designed for US populations, the 124-food item Dietary History Questionnaire (DHQ), was used to assess dietary intake, including frequency of consumption and portion size over the previous 12 months in the three cohorts.^23–25^ An earlier version of the 124-item DHQ was administered in the NIH-AARP from 1995-1996. The AHS DHQ was administered from 1999-2003 and the PLCO DHQ was administered from 1998-2000. The MyPyramid Equivalents Database, which is based on the US Department of Agriculture 1994-1996 Continuing Survey of Food Intakes by Individuals, was used to derive food group equivalents and energy intake estimates.^26^ Coffee intake (grams/day) was defined as the sum of regular and decaffeinated coffee. Tea intake (grams/day) was defined as the sum of regular and decaffeinated tea. Alcoholic beverage intake (grams/day) was defined as the sum of reported wine, beer, and liquor consumption and assumed gram weights of approximately 355g for a standard beer (12 fl oz), 148g for a standard pour of wine (5 fl oz), and 100-120g for a standard cocktail (1.5 fl oz), which will vary by mixer (USDA FoodData Central Database). SSB intake (grams/day) was defined as the sum of caffeinated and decaffeinated soda, caffeinated and decaffeinated sweet tea, and caffeinated and decaffeinated fruit punch (including lemonade, hi-C). ASB intake (grams/day) was defined as the sum of caffeinated and decaffeinated diet soda, caffeine and decaffeinated diet sweet tea, and caffeinated and decaffeinated diet fruit punch. SSBs and ASBs did not include hot coffee or hot tea with artificial sweeteners. Fruit juice intake (grams/day) was defined as reported 100% fruit juice consumption. All beverage variables were energy adjusted using the nutrient density method (grams/1000kcal/day). Coffee, tea, alcoholic beverages, SSBs, and ASBs were categorized as non-consumers compared to cohort and sex-specific tertiles (**Supplemental Table 1**). Fruit juice was categorized into cohort and sex-specific quartiles as most participants (93.7%, 5,243/5,598) consumed at least some amount of fruit juice.

### Demographic and lifestyle information

Demographic and lifestyle characteristics were collected using standardized questionnaires specific to each cohort and administered at baseline or during follow-up visits. Data related to age at oral sample collection, sex, self-reported race and ethnicity, education level, body mass index (BMI), smoking status, and total energy intake based on the lifestyle questionnaires or FFQs were included for analysis.

### Oral microbiome

DNA extraction, PCR amplification, sequencing, and bioinformatics procedures and methods have been previously described.^7^ Briefly, DNA was extracted from the buccal cell pellets using the DSP DNA Virus Pathogen kit (Qiagen). PCR amplification was conducted using barcoded primers targeting the 16S rRNA gene V4 region. Sequencing of the DNA was performed using the MiSeq platform (Illumina) with 2×250-bp paired-end reads. The sequencing data from the three cohorts were processed independently using QIIME 2 version 2018.4^27^ to produce amplicon sequence variants (ASVs)^28^ and taxonomy was assigned using the SILVA v132 database.^29^ Genus-level presence and relative abundances were estimated. Alpha diversity metrics (observed ASVs, Faith’s phylogenetic diversity (PD),^30^ and Shannon index^31^) and beta diversity metrics (Bray-Curtis dissimilarity, unweighted UniFrac, and weighted UniFrac^32^) were calculated based on even sampling depth of 20,000 reads per sample.

### Statistical analysis

For each cohort, demographic (age at oral sample collection, sex, education level, and self-identified race and ethnicity) and lifestyle (body mass index [BMI] and smoking status) characteristics, dietary factors, and alpha diversity indices were summarized in Table 1, adjusting for sample weights. All association analyses accounted for the complex sampling design using sampling weights and strata. Beta-diversity analyses were conducted separately within each cohort using Bray-Curtis, unweighted and weighted UniFrac distance matrices. We used *FastAdonis*, a computationally efficient PERMANOVA-based algorithm, accounting for sample weight, to estimate the R^2^, the proportion of beta-diversity variability explained by individual variables marginally and by the full covariate set jointly.^33^ Standard errors were obtained using weighted bootstrap resampling in *FastAdonis*. Cohort-specific R² estimates were combined using fixed-effect meta-analysis to obtain an overall R² estimate.

**Table 1.** Demographic, lifestyle, dietary, and oral microbiome characteristics of the participants in the Agricultural Health Study (AHS), NIH-AARP Diet and Health Study, and Prostate, Lung, Colorectal, and Ovarian Cancer.

|  | **AHS**<br>Unweighted N = 1,468<br>Weighted N = 26,752 | **NIH-AARP**<br>Unweighted N = 1,934<br>Weighted N = 25,084 | **PLCO**<br>Unweighted N = 2,196<br>Weighted N = 34,957 |
| --- | --- | --- | --- |
| **Age, years, mean (SD)** | 54.44 (11.57) | 70.99 (5.31) | 66.99 (5.72) |
| **Sex, n (%)** |  |  |  |
| Female | 565 (45.6) | 723 (40.1) | 1077 (54.0) |
| Male | 903 (55.4) | 1211 (59.9) | 1119 (46.0) |
| **BMI, kg/m2, n (%)** |  |  |  |
| <25 | 351 (26.5) | 728 (37.9) | 738 (33.0) |
| 25.0 to <30.0 | 431 (29.5) | 865 (44.1) | 980 (45.0) |
| ≥30 | 218 (12.7) | 341 (17.69) | 478 (22.0) |
| Missing | 468 (31.3) | 0 (0.0) | 0 (0.0) |
| **Education, n (%)** |  |  |  |
| <High school | 117 (6.3) | 93 (3.9) | 157 (4.9) |
| High school | 665 (40.7) | 316 (15.8) | 523 (21.3) |
| Some college | 347 (26.5) | 658 (33.2) | 778 (34.6) |
| College graduate | 259 (21.0) | 867 (47.1) | 738 (39.1) |
| Missing | 80 (5.5) | 0 (0.0) | 0 (0.0) |
| **Race and ethnicity, n (%)** |  |  |  |
| Non-Hispanic White | 1434 (98.0) | 1823 (94.0) | 2068 (94.3) |
| Other races and ethnicities | 34 (2.0) | 111 (6.0) | 128 (5.7) |
| **Smoking status, n (%)** |  |  |  |
| Current smoker | 330 (9.6) | 463 (9.7) | 616 (9.2) |
| Former smoker | 509 (26.1) | 1031 (48.9) | 932 (42.9) |
| Never smoker | 545 (57.4) | 355 (36.0) | 586 (43.2) |
| Never smoker but used cigars | 48 (4.8) | 31 (2.6) | 62 (4.7) |
| Missing | 36 (2.1) | 54 (2.8) | 0 (0.0) |
| **Total energy, kcal/day, mean (SD)** | 1969.44 (802.99) | 1810.87 (719.35) | 1723.32 (715.05) |
| **Beverages, g/day, mean (SD)** |  |  |  |
| Coffee | 636.91 (774.45) | 780.98 (642.42) | 874.98 (850.21) |
| Tea | 312.32 (533.82) | 207.13 (386.52) | 263.56 (475.72) |
| Alcohol | 72.51 (209.38) | 184.07 (606.25) | 129.00 (528.02) |
| SSBs | 200.48 (330.41) | 142.77 (341.37) | 92.23 (209.78) |
| ASBs | 210.84 (448.81) | 221.26 (441.81) | 162.89 (366.19) |
| Fruit juice | 118.85 (148.30) | 185.22 (251.79) | 137.17 (172.79) |
| **Alpha diversity, mean (SD)** |  |  |  |
| Observed ASVs | 124.57 (44.65) | 129.04 (42.10) | 122.87 (38.78) |
| Faith's PD | 10.33 (2.47) | 10.57 (2.37) | 9.21 (1.94) |
| Shannon Index | 4.35 (0.74) | 4.58 (0.65) | 4.46 (0.63) |

Note: Table counts are unweighted, but all percentages, means, and standard deviations (SD) are weighted estimates
Abbreviations: Amplicon sequence variants (ASVs), phylogenetic diversity (PD), saturated fatty acids (SFAs), unsaturated fatty acids (UFAs)

Although beverage intakes were measured continuously in grams per day and energy-adjusted using the nutrient density method (grams/1000 kcal/day), they were analyzed as categorical variables in the alpha-diversity and genus-level analyses. Trend tests were performed by assigning each intake category its cohort– and sex-specific median value and modeling this valuing as a continuous variable using the corresponding regression model; p-values were generated from the Wald test.

Associations of individual beverages with three alpha diversity metrics (observed ASVs, Faith’s phylogenetic diversity, and Shannon index) were evaluated separately within each cohort using weighted multivariable linear regression. Models were adjusted for demographic and lifestyle characteristics and were fit using the svyglm function in the R package survey. Cohort-specific estimates across cohorts were combined using fixed-effect meta-analysis to estimate pooled regression coefficients, 95% confidence intervals, and nominal p-values. Between-cohort heterogeneity was assessed using Cochran’s Q test.

For genus-level analyses, genus prevalence and sampling-weighted mean relative abundance were estimated across all study populations combined. Based on these pooled estimates, each genus was assigned to one of three mutually exclusive model categories for all beverage analyses. This categorization matched the model to the genus prevalence and relative abundance while avoiding testing the same beverage-genus association using multiple model types. First, genera with a prevalence of 5%– 95% and sampling-weighted mean relative abundance greater than 0.1% were analyzed using survey-weighted zero-inflated negative binomial (ZINB) regression models (n=21). The ZINB model provided two component-specific p-values, one for the zero-inflation component and one for the count component. For interpretability, odds ratios from the zero-inflation component were reciprocated so that values greater than 1 indicated greater odds of membership in the count-generating class.

Second, genera with a prevalence of 5%–95% and mean relative abundance less than or equal to 0.1% were analyzed using survey-weighted logistic regression models for genus presence versus absence (n=60, because presence was considered more reliably estimable than relative abundance for these low abundance taxa. Third, genera with a prevalence greater than 95% were analyzed using survey-weighted linear regression models of centered log-ratio (CLR) transformed relative abundance (n = 20 genera). A pseudocount of 1 × 10 was added to relative abundance values prior to CLR transformation. All regressions were adjusted for the same set of covariates as the alpha-diversity models. Cohort-specific genus-level estimates were combined using fixed-effect meta-analysis.

For each beverage, multiple-testing correction was performed using the Benjamini-Hochberg procedure by controlling for FDR at 0.05. Presence-based p-values from logistic regression models were corrected together with the zero-inflation-component p-values from ZINB models. Abundance-based p-values from CLR-linear models were corrected together with the count-component p-values from ZINB models. Thus, ZINB component p-values were adjusted within the corresponding testing family, and no genus was counted more than once within a given correction set. Any deviations for heterogeneity were noted. All statistical analyses were performed using R version 4.5.1, except survey-weighted ZINB models, which were fit using Stata/SE 19.

## Results

### Population characteristics

Participants included in this study had a mean age of 54.4 years (SD 11.6) in AHS, 71.0 years (SD 5.3) in NIH-AARP, and 67.0 years (SD 5.7) in PLCO at sample collection (**Table 1**). In general, the proportions of men and women were similar across the three cohorts, and all cohorts were composed of pre-dominantly non-Hispanic White individuals. AHS had the highest average reported consumption of tea and SSBs and the lowest average reported consumption of alcoholic beverages. NIH-AARP had the highest average reported consumption of alcoholic beverages, ASBs, and fruit juice, and PLCO had the highest average reported consumption of coffee. Overall, associations were generally consistent across cohorts, with little evidence of between-cohort heterogeneity.

### Variability in beta diversity matrices explained by beverages and other factors

For the three cohorts combined, a joint model containing all beverages and baseline factors explained 7.50% of the variability in unweighted-UniFrac, 8.30% in weighted UniFrac and 7.53% in Bray-Curtis, while a model containing all beverages jointly explained 4.06%, 4.94%, and 4.21% of the variability in these beta diversity indices, respectively (**Supplementary Figure 1**).

In separate univariable models of individual beverages and baseline factors, the largest proportion of variability in the unweighted-UniFrac distance matrix was explained by smoking status (1.23%), followed by age (0.99%), education (0.89%), and alcoholic beverage intake (0.86%) (**Figure 1**). Total alcoholic beverage intake explained more variability than any individual alcohol type. Among the individual alcohol types, beer explained the greatest proportion of variability in unweighted UniFrac (0.81%), whereas wine and liquor explained 0.60% and 0.59%, respectively.

**Figure 1.**
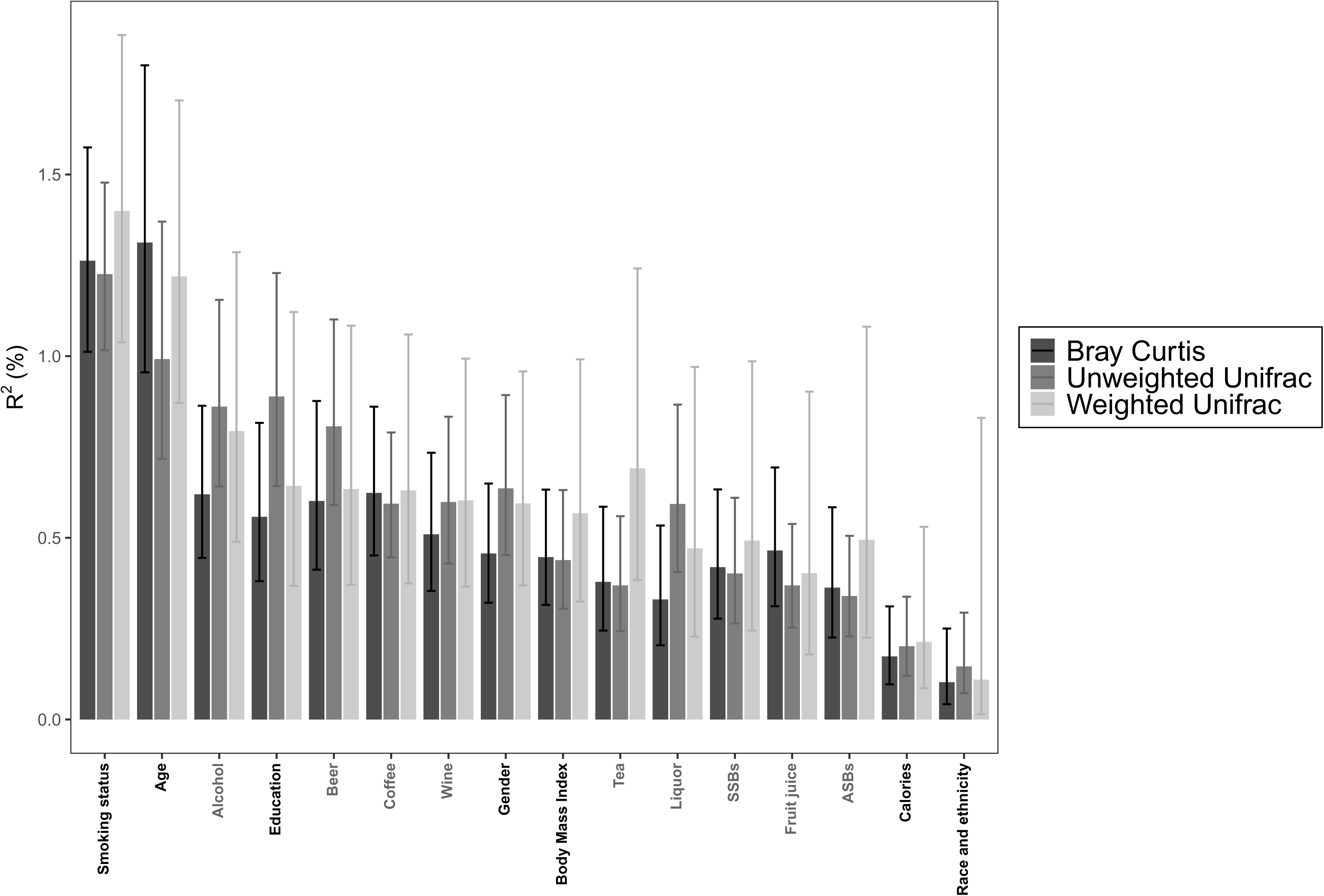
Variability in Bray-Curtis, unweighted UniFrac, and weighted UniFrac beta diversity matrices explained by demographic, lifestyle, and beverage variables. The FastAdonis function in R was used to calculate the variability explained by these variables.

### Beverage associations with alpha diversity

Alcoholic beverage intake was positively associated with alpha diversity, with 10.18 more observed ASVs among participants in the highest tertile of alcoholic beverage intake compared with non-consumers (95% CI _T3 vs. non-consumers_: 5.56, 14.80, p-trend=0.0002) (**Table 2**, see **Supplemental Table 2** for individual cohorts). Both the Shannon index (β_T3 vs. non-consumers_: 0.12 [95% CI: 0.04, 0.19], p-trend<0.0001) and Faith’s PD (β_T3 vs. non-consumers_: 0.62 [95% CI: 0.37, 0.86], p-trend < 0.0001) had similar trends with alcoholic beverages (**Table 2**). When further categorized by type of alcoholic beverage consumption (i.e., beer, wine, liquor), each type was associated with higher observed ASVs and Faith’s PD, however, only liquor was associated with higher Shannon index (**Table 2**). For example, the highest tertile of liquor consumption was associated with 15.21 additional ASVs compared to non-consumers (95% CI: 7.91, 22.50, p-trend=0.0003), while beer and wine were respectively associated with an additional 6.72 (95% CI: 1.93, 11.51, p-trend = 0.0314) and 7.04 (95% CI: 2.32, 11.77, p-trend =0.0226) more observed ASVs compared to non-consumers.

**Table 2.** Meta analysis of linear regression associations between coffee, tea, alcohol, SSBs, ASBs, and fruit juice with alpha diversity metrics.

|  | Observed ASVs |  |  |  | Shannon index |  |  |  | Faith's phylogenetic diversity |  |  |  |
| --- | --- | --- | --- | --- | --- | --- | --- | --- | --- | --- | --- | --- |
|  | Beta (95% CI) | p-value | p-trend | p-hetero | Beta (95% CI) | p-value | p-trend | p-hetero | Beta (95% CI) | p-value | p-trend | p-hetero |
| **Coffee** |  |  |  |  |  |  |  |  |  |  |  |  |
| T1 vs. non-consumers | -3.84 (-8.61, 0.93) | 0.1149 |  | 0.6779 | -0.07 (-0.16, 0.01) | 0.0842 |  | 0.3352 | -0.11 (-0.36, 0.14) | 0.384 |  | 0.7853 |
| T2 vs. non-consumers | -0.33 (-5.32, 4.66) | 0.8958 | 0.2764 | 0.2798 | -0.03 (-0.12, 0.05) | 0.4319 | 0.4749 | 0.0547 | 0.04 (-0.22, 0.30) | 0.7575 | 0.1942 | 0.3399 |
| T3 vs. non-consumers | 0.86 (-4.88, 6.61) | 0.7685 |  | 0.0889 | -0.01 (-0.10, 0.09) | 0.9063 |  | 0.0460 | 0.13 (-0.16, 0.43) | 0.3756 |  | 0.0603 |
| **Tea** |  |  |  |  |  |  |  |  |  |  |  |  |
| T1 vs. non-consumers | 2.97 (-1.41, 7.34) | 0.1837 |  | 0.835 | 0.02 (-0.05, 0.09) | 0.5773 |  | 0.7387 | 0.18 (-0.05, 0.42) | 0.1222 |  | 0.9791 |
| T2 vs. non-consumers | -1.40 (-6.00, 3.21) | 0.5523 | 0.7452 | 0.4409 | -0.04 (-0.12, 0.03) | 0.254 | 0.9828 | 0.2029 | -0.03 (-0.27, 0.21) | 0.8044 | 0.7782 | 0.6702 |
| T3 vs. non-consumers | 1.09 (-3.36, 5.54) | 0.6301 |  | 0.5281 | 0.00 (-0.07, 0.08) | 0.9007 |  | 0.0564 | 0.07 (-0.16, 0.31) | 0.5526 |  | 0.7175 |
| **Alcohol (all)** |  |  |  |  |  |  |  |  |  |  |  |  |
| T1 vs. non-consumers | 3.61 (-0.77, 7.99) | 0.1059 |  | 0.6698 | 0.05 (-0.02, 0.12) | 0.1603 |  | 0.0505 | 0.22 (-0.01, 0.45) | 0.0565 |  | 0.5978 |
| T2 vs. non-consumers | 5.88 (1.34, 10.43) | 0.0112 | 0.0002 | 0.8555 | 0.04 (-0.04, 0.11) | 0.3184 | <0.0001 | 0.4758 | 0.31 (0.07, 0.55) | 0.0102 | <0.0001 | 0.8053 |
| T3 vs. non-consumers | 10.18 (5.56, 14.80) | <0.0001 |  | 0.5186 | 0.12 (0.04, 0.19) | 0.0016 |  | 0.6677 | 0.62 (0.37, 0.86) | <0.0001 |  | 0.3172 |
| **Beer** |  |  |  |  |  |  |  |  |  |  |  |  |
| T1 vs. non-consumers | 3.80 (-0.59, 8.19) | 0.0896 |  | 0.5437 | 0.05 (-0.02, 0.12) | 0.1818 |  | 0.1195 | 0.22 (-0.01, 0.46) | 0.0625 |  | 0.7636 |
| T2 vs. non-consumers | 6.11 (1.58, 10.64) | 0.0082 | 0.0314 | 0.4952 | 0.09 (0.02, 0.16) | 0.0164 | 0.0485 | 0.1076 | 0.29 (0.06, 0.53) | 0.0149 | 0.0046 | 0.7469 |
| T3 vs. non-consumers | 6.72 (1.93, 11.51) | 0.006 |  | 0.6838 | 0.05 (-0.03, 0.13) | 0.1964 |  | 0.7073 | 0.44 (0.18, 0.70) | 0.0008 |  | 0.3621 |
| **Wine** |  |  |  |  |  |  |  |  |  |  |  |  |
| T1 vs. non-consumers | 6.14 (2.06, 10.21) | 0.0032 |  | 0.9091 | 0.04 (-0.02, 0.11) | 0.2071 |  | 0.3154 | 0.29 (0.08, 0.51) | 0.0067 |  | 0.8572 |
| T2 vs. non-consumers | 6.01 (1.52, 10.49) | 0.0087 | 0.0226 | 0.9162 | 0.05 (-0.02, 0.13) | 0.1825 | 0.0558 | 0.5016 | 0.35 (0.11, 0.58) | 0.0036 | 0.0131 | 0.9031 |
| T3 vs. non-consumers | 7.04 (2.32, 11.77) | 0.0035 |  | 0.4276 | 0.06 (-0.02, 0.13) | 0.1436 |  | 0.1224 | 0.40 (0.15, 0.65) | 0.0021 |  | 0.4290 |
| **Liquor** |  |  |  |  |  |  |  |  |  |  |  |  |
| T1 vs. non-consumers | 6.48 (3.07, 9.90) | 0.0002 |  | 0.8897 | 0.08 (0.03, 0.14) | 0.0032 |  | 0.7223 | 0.33 (0.15, 0.51) | 0.0003 |  | 0.8115 |
| T2 vs. non-consumers | 7.37 (1.73, 13.01) | 0.0104 | 0.0003 | 0.2976 | 0.12 (0.02, 0.21) | 0.0145 | 0.0003 | 0.0431 | 0.48 (0.16, 0.79) | 0.0036 | <0.0001 | 0.4565 |
| T3 vs. non-consumers | 15.21 (7.91, 22.50) | <0.0001 |  | 0.4561 | 0.21 (0.09, 0.34) | 0.0006 |  | 0.9034 | 0.83 (0.45, 1.21) | <0.0001 |  | 0.6410 |
| **SSBs** |  |  |  |  |  |  |  |  |  |  |  |  |
| T1 vs. non-consumers | 0.98 (-3.49, 5.45) | 0.6676 |  | 0.6686 | 0.05 (-0.02, 0.12) | 0.1732 |  | 0.7363 | 0.05 (-0.19, 0.28) | 0.6852 |  | 0.6667 |
| T2 vs. non-consumers | 3.37 (-1.16, 7.89) | 0.1453 | 0.5458 | 0.6965 | 0.05 (-0.02, 0.13) | 0.1519 | 0.6348 | 0.5857 | 0.10 (-0.14, 0.33) | 0.4269 | 0.1224 | 0.4455 |
| T3 vs. non-consumers | -2.45 (-7.06, 2.16) | 0.2977 |  | 0.3129 | 0.01 (-0.07, 0.08) | 0.8044 |  | 0.6735 | -0.24 (-0.48, 0.00) | 0.0493 |  | 0.2454 |
| **ASBs** |  |  |  |  |  |  |  |  |  |  |  |  |
| T1 vs. non-consumers | 1.67 (-2.87, 6.20) | 0.471 |  | 0.9158 | 0.02 (-0.05, 0.09) | 0.6074 |  | 0.4358 | 0.12 (-0.12, 0.36) | 0.3161 |  | 0.8553 |
| T2 vs. non-consumers | -0.21 (-4.32, 3.90) | 0.9205 | 0.1409 | 0.7119 | -0.03 (-0.10, 0.03) | 0.339 | 0.8729 | 0.5743 | 0.03 (-0.19, 0.24) | 0.8083 | 0.2495 | 0.5024 |
| T3 vs. non-consumers | -2.76 (-7.21, 1.69) | 0.2236 |  | 0.8449 | -0.01 (-0.08, 0.07) | 0.8795 |  | 0.1620 | -0.10 (-0.34, 0.14) | 0.3968 |  | 0.7245 |
| **Fruit juice** |  |  |  |  |  |  |  |  |  |  |  |  |
| Q2 vs. Q1 | 0.85 (-3.65, 5.35) | 0.7118 |  | 0.2601 | 0.06 (-0.02, 0.13) | 0.128 |  | 0.2722 | 0.06 (-0.18, 0.30) | 0.6158 |  | 0.5575 |
| Q3 vs. Q1 | 1.72 (-2.76, 6.19) | 0.4523 | 0.8593 | 0.5616 | 0.01 (-0.06, 0.09) | 0.7044 | 0.6819 | 0.6797 | 0.12 (-0.12, 0.36) | 0.3193 | 0.6536 | 0.7072 |
| Q4 vs. Q1 | -0.91 (-5.34, 3.52) | 0.6879 |  | 0.824 | 0.03 (-0.04, 0.10) | 0.4599 |  | 0.9037 | -0.07 (-0.31, 0.16) | 0.5404 |  | 0.5379 |

SSBs showed a modest inverse association with Faith’s PD (β_T3 vs. non-consumers_: –0.24 [95% CI: – 0.48, 0.00], p-trend = 0.1224); however, the trend was not statistically significant. SSBs were not statistically significantly associated with observed ASVs (β_T3 vs. non-consumers_: –2.45 [95% CI: –7.06, 2.16], p-trend = 0.5458) or Shannon index (β_T3 vs. non-consumers_: 0.01 [95% CI: –0.07, 0.08], p-trend = 0.6348). Coffee, tea, ASBs, and fruit juice were not associated with any alpha diversity measures.

### Beverage associations with genus-level prevalence and relative abundance

At the genus level, most of the observed significant associations (i.e., FDR < 0.05) were with alcoholic beverages; few associations were seen for coffee, and no association was observed for tea, SSBs, ASBs, or fruit juice (**Figure 2**). In the highest tertile of alcoholic beverage consumption, the minimum g/day value was 30.1 g/day; this was for females in AHS. The maximum g/day value for the highest tertile was 9870.5 g/day and was observed for males in PLCO (**Supplemental Table 1**). We observed significant associations with the highest tertile of alcoholic beverage consumption compared to non-consumption for seven genera for the zero-inflation component and one genus for the negative-binomial component of the ZINB models, 15 genera for the logistic regression models, and two genera for the CLR-transformed linear regression models.

**Figure 2.**
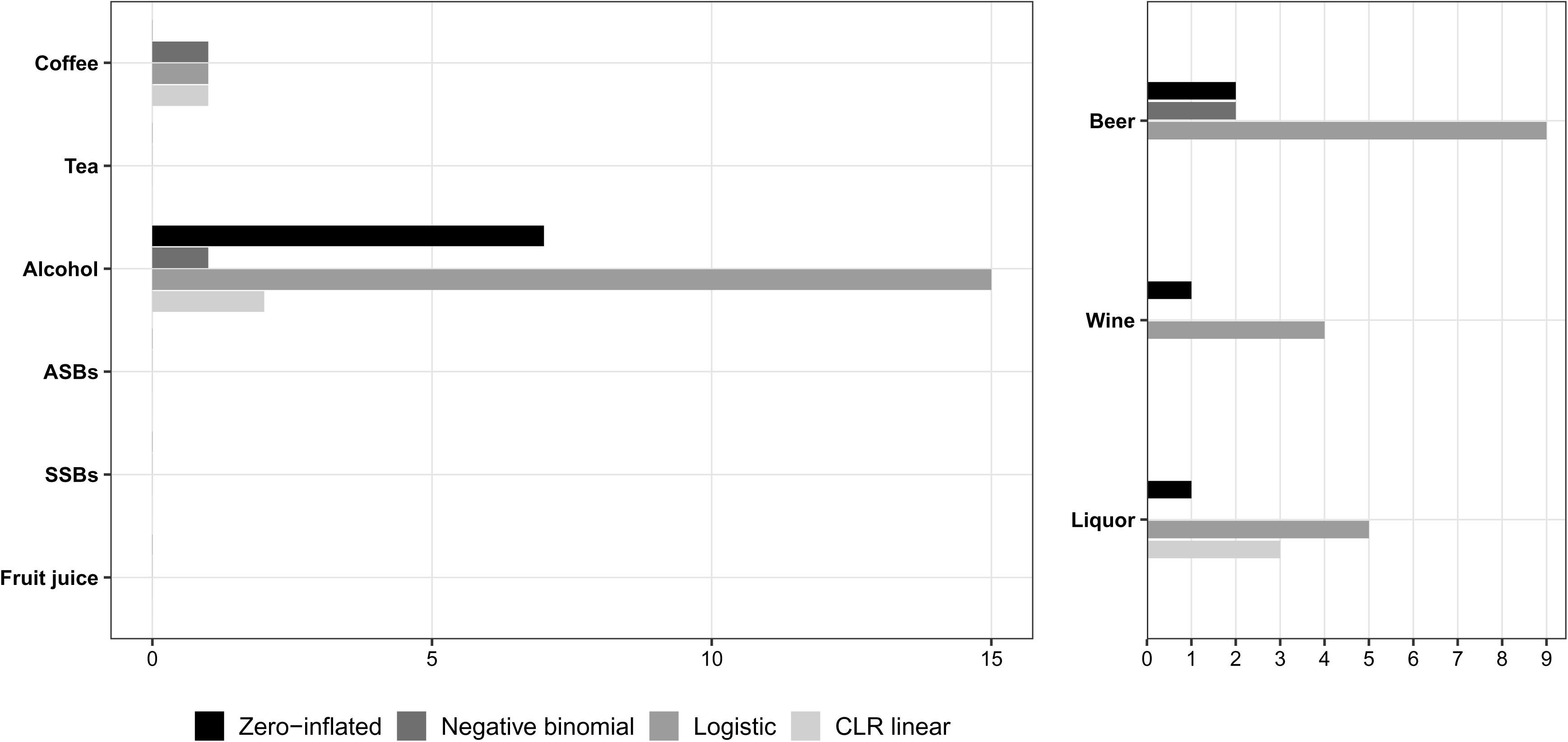
Number of genus-level associations for each beverage and beverage type (for alcohol only) by regression model type.

For overall alcoholic beverage intake, in the ZINB models (used when genera were present in 5-95% of participants and had a weighted relative abundance greater than 0.1% [n=21 genera]), the reported ORs from the zero-inflation component were 2.25 for *Megasphaera* (95% CI: 1.43, 3.54; p-trend=0.0157), 2.17 for *Peptostreptococcus* (95% CI: 1.63, 2.89; p-trend=0.0012), and 1.95 for *Moryella* (95% CI: 1.51, 2.52; p-trend=0.0001) for the highest tertile compared with non-consumers. A similar component-specific association for *Peptostreptococcus* was observed for beer (OR_T3 vs. non-consumers_: 2.03 [95% CI: 1.48, 2.78], p-trend = 0.0200) and liquor (OR_T3 vs. non-consumers_: 1.76 [95% CI: 1.21, 2.56], p-trend = 0.3354), whereas the association with *Moryella* was observed for liquor (OR_T3 vs. non-consumers_: 1.67 [95% CI: 1.19, 2.34], p-trend=0.0344) and wine (OR_T3 vs. non-consumers_: 1.91 [95% CI: 1.47, 2.49], p-trend=0.0027). The zero-inflation component also showed an association between alcoholic beverage consumption and *Eubacterium nodatum* group (OR_T3 vs. non-consumers_: 1.79 [95% CI: 1.32, 2.43], p-trend = 0.0190). For alcohol overall, an inverse zero-inflated component-specific association was observed for *Scardovia* (OR_T3 vs. non-consumers_: 0.65 [95% CI: 0.49, 0.86], p-trend = 0.0050). For beer, the negative binomial component for *Scardovia* yielded a RR of 0.53 (95% CI: 0.39, 0.74, p-trend = 0.008), and the zero-inflated component yielded an OR of 0.63 (95% CI: 0.47, 0.85, p-trend = 0.0658). Additionally, in the negative-binomial component, the highest tertile of alcoholic beverage consumption was associated with a 23% lower expected *Lautropia* abundance compared with non-consumers (RR_T3 vs. non-consumers_: 0.77 [95% CI: 0.65, 0.91], p-trend = 0.0045); a very similar association was observed for beer, but not the other alcohol types (**Supplemental Table 3).**

In the logistic regression models (used when genera were present in 5-95% of the participants with weighted relative abundance less than or equal to 0.1% [n=60 genera]), for the highest consumers of alcoholic beverages, the odds of detectable Firmicutes oral clone FM046 and Defluviitaleaceae UCG-011 were 2.87 (95% CI: 1.83, 4.50, p-trend = 0.0005) and 2.19 (95% CI: 1.59, 3.01, p-trend <0.0001) times that of non-consumers, respectively. When further examining alcoholic beverage types, the relationship with Firmicutes oral clone FM046 was strongest for liquor (OR_T3 vs. non-consumers_: 4.41 [95% CI: 2.45, 7.94], p-trend <0.0001) and the association with Defluviitaleaceae UCG-011 was only observed for beer (OR_T3 vs. non-consumers_: 2.07 [95% CI: 1.51, 2.83], p-trend = 0.0136). Further, higher alcoholic beverage intake was associated with higher odds of the presence of *Filifactor* (OR_T3 vs. non-consumers_: 1.71 [95% CI: 1.30, 2.26], p-trend = 0.0022), particularly for beer drinkers (OR_T3 vs. non-consumers_: 1.73 [95% CI: 1.31, 2.27], p-trend = 0.0030), and *Butyrivibrio 2* (OR_T3 vs. non-consumers_: 1.53 [95% CI: 1.20, 1.94], p-trend <0.0001), particularly for wine (OR_T3 vs. non-consumers_: 1.73 [95% CI: 1.35, 2.23], p-trend = 0.0002) or liquor drinkers (OR_T3 vs. non-consumers_: 1.80 [95% CI: 1.20, 2.68], p-trend = 0.0012). Among genera evaluated using logistic regression, the only inverse association between alcoholic beverage intake and the presence of genus-level bacteria was for *Bifidobacterium* (OR_T3 vs. non-consumers_: 0.67 [95% CI: 0.53, 0.86], p-trend = 0.0084), however it was associated only with alcoholic beverages overall and not specific types of alcoholic beverages (**Supplemental Table 4**).

For the CLR-transformed linear regression models (used for genera present in more than 95% of participants [n=20 genera]), the CLR-transformed abundance of *Alloprevotella* was higher for alcoholic beverage intake overall (β_T3 vs. non-consumers_: 0.16 [95% CI: 0.05 0.26], p-trend = 0.0014) and the CLR-transformed abundance of *Streptococcus* was lower in the highest tertile of alcohol drinkers (β_T3 vs. non-consumers_: –0.21 [95% CI: –0.32, –0.10], p-trend = 0.0002), particularly those in the highest tertile of liquor drinkers (β_T3 vs. non-consumers_: –0.31 [95% CI: –0.48, –0.14], p-trend = 0.0268). (**Supplemental Table 5**).

Three of the 28 total significant beverage associations were with coffee (**Supplemental Tables 3-5**). For example, in the linear regression models of CLR-transformed abundances, the highest tertile of coffee intake was associated with higher CLR-transformed abundance of *Atopobium* compared with non-consumers (β_T3 vs. non-consumers_: 0.25 [95% CI: 0.11, 0.38], p-trend = 0.0021 **(Supplemental Tables 3-5).**

## Discussion

This cross-sectional study is the largest to date investigating associations between the consumption of different beverages and the oral microbiome. Of all beverages examined, alcohol appeared to be most strongly related to measures of the oral microbiome. Greater alcohol consumption was associated with higher alpha diversity estimates and multiple genus-level measures, including associations involving *Peptostreptococcus, Moryella, Megasphaera,* and *Scardovia*. We identified some associations with coffee, particularly with beta-diversity variability and some genera. SSBs showed little association with alpha diversity or beta-diversity variability and no FDR-significant genus-level associations. Tea, ASBs, and fruit juice consumption showed no notable associations with the oral microbiome measures examined.

Previous studies have consistently identified associations between alcohol and the overall microbial community with alcohol as a top contributor to beta diversity, but inconsistent alpha diversity and taxa-level associations have been observed.^17–19,34,35^ With respect to alpha diversity, consistent with our finding that overall alcohol consumption was associated with higher alpha diversity and that each specific alcohol type was associated with higher observed ASVs and Faith’s PD, two epidemiologic studies, a cohort of postmenopausal women in the Osteoporosis and Periodontal Disease Study which is an ancillary study of the Women’s Health Initiative (WHI), and a cross-sectional study set in two U.S. cohorts (CPS-II Nutrition cohort and the PLCO cohort) found that moderate to heavy drinkers or participants in the highest alcohol intake group had greater alpha diversity compared to non-drinkers.^17,18^ In another study, a cross-sectional analysis of heavy compared to light drinkers, no associations were observed for alpha diversity.^34^ Conversely, a cross-sectional study in Italy observed a lower species richness for red wine drinkers compared to a reference group of water drinkers; however, this association was determined through targeted PCR-based methods rather than more comprehensive methods such as sequencing.^35^ The ancillary study from WHI, which also investigated individual types of alcohol beyond general alcohol consumption, found that the association with increased alpha diversity remained for wine, but not beer or liquor.^17^ Prior literature suggests that the presence of polyphenolic compounds in wine may suppress bacterial growth by inhibiting the formation of bacterial biofilm; this represents an area to further explore with mechanistic research.^36^

At the genus level, associations observed in our study included an alcohol-related association involving *Peptostreptococcus*, a genus associated with periodontal disease,^37^ as well as an inverse association with *Lautropia*, a commensal bacterium often at lower abundance in dental diseases^38^ and *Streptococcus*. These associations were not reported in prior studies. One prior study reported positive associations between alcohol and Lactobacillales and Bacilli,^18^ whereas we observed an inverse association between alcohol, particularly liquor, and *Streptococcus*, a member of the Lactobacillales order. The inverse relationship between alcohol consumption and *Streptococcus* is of interest given that several *Streptococcus* species are capable of converting ethanol to acetaldehyde.^39,40^ Consistent with our findings, the ancillary study from WHI observed lower abundance of *Streptococcus mutans* with increased alcohol consumption, although this association was not statistically significant after multiple testing correction. While the cross-sectional nature of these data precludes the assessment of temporality, these findings motivate further investigation of a potential relationship between alcohol consumption and the functional capacity of the oral microbiome. Most alcohol-associated genera were identified either through the zero-inflation component of the ZINB models or through logistic regression models of observed genus presence. Together with the relatively greater contribution of alcohol to variability in the presence/absence-based unweighted UniFrac distance, this pattern suggests that alcohol may be more strongly related to occurrence patterns of specific oral taxa than to shifts in relative abundance among highly prevalent taxa. However, for the associations of alcohol types (wine, beer, and liquor) no information regarding specific wine, beers, or liquor and mixers were assessed, which may limit the associations observed.

Although coffee and wine both contain polyphenolic compounds^36^, few associations were observed for coffee in our analysis. We observed no associations for coffee with any alpha diversity measures, minimal genus-level associations, and in univariable analysis, coffee explained a smaller percent of variability (0.59%, 0.63%) in the overall microbial community than alcohol (0.86%, 0.79%) for unweighted and weighted UniFrac distances, respectively. The cross-sectional study in Italy mentioned previously also examined coffee intake,^35^and in contrast to our study, coffee intake was associated with greater species richness. Additionally, habitual coffee intake was associated with decreased detection of periodontal pathogens compared to habitual water drinkers.^35^ In the prior CPS-II and PLCO cross-sectional analyses, coffee was not associated with any alpha or beta diversity metrics, but was positively associated with *Granulicatella* and *Synergistetes* abundances, in contrast to our results.^16^

For tea, SSBs, ASBs, and fruit juice, fewer studies of associations with the oral microbiome have been conducted. In the same prior analyses of CPS-II and PLCO cohorts, there was a positive association of tea with alpha diversity measures, shifts in beta diversity (weighted UniFrac), and a positive association with *Clostridia*, Fusobacteriales, and lower abundances of Lactobacillales and *Bifidobacteriaceae*,^16^ which contrasts with our study where no associations between tea consumption and the oral microbiome were observed. Moreover, we observed no significant associations of SSBs with alpha diversity and no significant genus-level associations, while the previous study in the CPS-II and the PLCO cohorts found lower richness and differences in beta diversity for those with higher compared to lower SSB intake.^14^ Higher SSB intake was also associated with greater relative abundances of *Bifidobacteriaceae*, *Lactobacillus rhamonosus*, and *Streptococcus tigurinus*, and lower relative abundances of *Lachnoanaerobaculum*, Fusobacteriales/*Leptotrichia*, and *Campylobacter* in this study.^14^ In our study, there was a greater proportion of non-consumers of SSBs (1747/5598, 31.2%) and ASBs (2550/5598, 45.6%) than of tea (1365/5598, 24.4%), coffee (762/5598, 13.6%), and alcohol (1486/5598, 26.5%). Differences in these exposure distributions may contribute to variation in observed associations across beverage types and limit comparability with findings from studies with different consumption patterns.

Regarding fruit juice and ASBs, the existing evidence is mainly derived from intervention trials. A three-day, randomized intervention study compared three diets: juice only, juice and ad libitum diet, and a plant-based whole-food diet; they reported increases in the relative abundance of pro-inflammatory bacteria (e.g., Proteobacteria) at the family level in the oral microbiome for all diets over the course of the study, but the magnitude of the increase was the largest for the exclusive juice diet.^41^ However, the study included only 14 participants across three diet groups, limiting the statistical power.^41^ Randomized studies of sweetener exposures have also reported changes in the oral microbiome.^42,43^ These include a single-center, double blind, parallel randomized trial of a mouth rinse containing nutritive sweetener,^42^ and a larger randomized study of adults who were randomized to different types of non-nutritive sweeteners (e.g., sucralose, aspartame, stevia, saccharin)^43^, observed modulations to the oral microbiome. In the mouth-rinse study, inulin was associated with decreased alpha diversity and increased *Fusobacterium periodonticum* over a two-week period were observed in the study with the mouth rinse and non-nutritive sweetener.^42^ In the larger randomized study with different types of non-nutritive sweeteners, sucralose was associated with multiple *Streptococcus* species, saccharin with decreased *Fusobacterium*, aspartame with decreased *Porphyromonas* and *Prevotella* species, and no changes among controls.^43^

There are several limitations of this study. First, because this analysis of the case-cohort data was cross-sectional, temporality cannot be established for the relationship between beverage consumption and the oral microbiome. Further, dietary assessment and oral wash sample collection were not contemporaneous in all cohorts, and the interval between them varied across cohorts. The limited between-cohort heterogeneity nevertheless supports the consistency of the observed associations across these three study populations. Second, although dietary data were collected via a validated instrument, FFQs inherently have measurement error, which may have resulted in misclassification of the beverage consumption estimates. However, regularly consumed beverages such as coffee and tea are correlated across serial questionnaires, even with time in between; beverages consumed more episodically versus habitually would therefore likely have more inherent measurement error.^44^ FFQs also provide average or habitual dietary intake, rather than recent intake; therefore, our findings reflect associations with habitual beverage intake rather than short-term intake near the time of oral sample collection.

We measured the oral microbial communities using 16S rRNA gene sequencing, which generally supports taxonomic characterization at the genus level, therefore, future studies should use techniques such as shotgun metagenomic sequencing to obtain greater species-level resolution and information on microbial gene and functional potential. Lastly, a prior analysis of the U.S. population oral microbiome in NHANES identified oral hygiene practices as an important source of variability in oral microbiome composition.^19^ Lack of information on oral hygiene practices, along with factors such as medication use, may therefore have resulted in residual confounding.

This study also has several strengths that increase the rigor of the findings. This is, to our knowledge, the largest study to date to evaluate the association between the consumption of multiple beverages and the oral microbiome, providing increased statistical power to detect associations. In addition, the inclusion of three large, distinct, US cohorts allowed us to evaluate the consistency of beverage-oral microbiome associations across different study populations.

## Conclusion

We evaluated the relationship between different beverages and the oral microbiome in three large US cohort studies. Coffee was associated with a few oral bacteria, whereas alcohol was associated with multiple measures of the oral microbiome including increases in alpha diversity and presence of bacteria often associated with poor oral health, and the absence of bacteria associated with better oral health. Of the alcohol types, most associations were with beer or liquor, not wine. These findings underscore a potential oral microbiome-mediated pathway for the relationship between alcohol and systemic disease risk. Longitudinal studies are needed to confirm causality of relationships and future research is needed to understand the biochemical actions of specific components within the different types of alcohol to further elucidate the mechanisms related to oral health and systemic disease.

## Supporting information

NIH Cover Sheet

## Acknowledgements

The authors acknowledge the research contributions of the Cancer Genomics Research Laboratory of the Division of Cancer Epidemiology & Genetics at the National Cancer Institute for their expertise, execution, and support of this research in the areas of project planning, wet laboratory processing of specimens, and bioinformatics analysis of generated data. We also acknowledge the contributions of Dr. Nathaniel Rothman for his role in initiating and designing the oral rinse collection in the Agricultural Health Study. The content of this publication does not necessarily reflect the views or policies of the Department of Health and Human Services, nor does mention of trade names, commercial products, or organizations imply endorsement by the US Government.

## Contributions

Conceptualization: CCA, JS, RS, EL, EV

Statistical analyses: LEF, SAS, YY, XH, YW, VCC, JS

Resources: CLD, KJ, BDH, AH, LML, WYH, NDF, LEBF, DPS, RS

Writing – original draft: LEF

Writing – review and editing: LEF, SAS, YY, XH, YW, VCC, CLD, KJ, BDH, AH, LML, WYH, NDF, LEBF, DPS, CCA, JS, RS, EL, and EV

Supervision: EV

Funding acquisition: CCA, RS

## Data availability statement

Microbiome sequencing data can be found at the Sequence Read Archive (SRA), project number PRJNA801882 with limited metadata. To access complete metadata, an application must be approved from the Agricultural Health Study (www.aghealthstars.com), the NIH-AARP Diet and Health Study (www.nihaarpstars.com), and the Prostate, Lung, Colorectal, and Ovarian Cancer Screening Trial (www.cdas.cancer.gov).

## Funding/author disclosures

This research was supported by the Intramural Research Program of the National Institutes of Health (NIH), the National Institute of Environmental Health Sciences (Z01 ES049030), the National Cancer Institute (NCI) (Z01 CP010119), and the NCI’s Cancer Prevention Fellowship Program. This project has been funded in whole or in part with Federal funds from the National Cancer Institute, National Institutes of Health, under Contract No. 75N91019D00024. The content of this publication does not necessarily reflect the views or policies of the Department of Health and Human Services, nor does the mention of trade names, commercial products, or organizations imply endorsement by the U.S. Government. No potential conflicts of interest were reported by any of the authors.

## Abbreviations

AHS: Agricultural Health Study
ASB: Artificially sweetened beverage
ASV: Amplicon sequence variant
BMI: Body mass index
CI: Confidence interval
CLR: Centered log ratio
CPS-II: Cancer Prevention Study II
DHQ: Diet history questionnaire
FFQ: Food frequency questionnaire
HEI: Healthy Eating Index
PD: Phylogenetic diversity
PERMANOVA: Permutation multivariate analysis of variance
PLCO: Prostate, Lung, Colorectal, and Ovarian Cancer Screening Trial
SD: Standard deviation
SSB: Sugar sweetened beverage

## Notes

### Competing Interest Statement

The authors have declared no competing interest.

### Author Declarations

IRB of the National Cancer Institute waived ethical approval for this work.

## References

1. Baker JL, Mark Welch JL, Kauffman KM, McLean JS, He X. The oral microbiome: diversity, biogeography and human health. Nat Rev Microbiol. 2024;22(2):89–104.

2. Siddiqui R, Badran Z, Boghossian A, Alharbi AM, Alfahemi H, Khan NA. The increasing importance of the oral microbiome in periodontal health and disease. Future Sci OA. 2023;9(8):Fso856.

3. Deo PN, Deshmukh R. Oral microbiome: Unveiling the fundamentals. J Oral Maxillofac Pathol. 2019;23(1):122–128.

4. Peng X, Cheng L, You Y, et al. Oral microbiota in human systematic diseases. Int J Oral Sci. 2022;14(1):14.

5. Kwak S, Wang C, Usyk M, et al. Oral Microbiome and Subsequent Risk of Head and Neck Squamous Cell Cancer. JAMA Oncol. 2024;10(11):1537–1547.

6. Meng Y, Wu F, Kwak S, et al. Oral Bacterial and Fungal Microbiome and Subsequent Risk for Pancreatic Cancer. JAMA Oncol. 2025.

7. Vogtmann E, Hua X, Yu G, et al. The Oral Microbiome and Lung Cancer Risk: An Analysis of 3 Prospective Cohort Studies. J Natl Cancer Inst. 2022;114(11):1501–1510.

8. Vogtmann E, Yano Y, Zouiouich S, et al. The human oral microbiome and risk of colorectal cancer within three prospective cohort studies in the United States. Cancer. 2025;131(6):e35802.

9. Sedghi L, DiMassa V, Harrington A, Lynch SV, Kapila YL. The oral microbiome: Role of key organisms and complex networks in oral health and disease. Periodontol 2000. 2021;87(1):107–131.

10. Mukherjee C, Moyer CO, Steinkamp HM, et al. Acquisition of oral microbiota is driven by environment, not host genetics. Microbiome. 2021;9(1):54.

11. Martin CC, JC; Moshfegh, AJ. Beverage Choices among Adults: What We Eat in America, NHANES 2017-2018. Food Surveys Research Group Data Brief No. 31. 2020.

12. Jamali M, Anvarifard P, Hosseini B, et al. Sweetened beverages and cardiovascular outcomes: an umbrella review of mortality and health outcomes. Nutr J. 2025;24(1):173.

13. Shroff P, Gondivkar SM, Kumbhare SP, Sarode S, Gadbail AR, Patil S. Analyses of the Erosive Potential of Various Soft Drinks and Packaged Fruit Juices on Teeth. J Contemp Dent Pract. 2018;19(12):1546–1551.

14. Fan X, Monson KR, Peters BA, et al. Altered salivary microbiota associated with high-sugar beverage consumption. Sci Rep. 2024;14(1):13386.

15. Jin B, Chen H, Liu P, et al. Assessing the association between tea intake and risk of dental caries and periodontitis: a two-sample Mendelian randomization study. Sci Rep. 2024;14(1):4728.

16. Peters BA, McCullough ML, Purdue MP, et al. Association of Coffee and Tea Intake with the Oral Microbiome: Results from a Large Cross-Sectional Study. Cancer Epidemiol Biomarkers Prev. 2018;27(7):814–821.

17. Maley SJ, Yue Y, Burns KF, et al. Alcohol Consumption and the Diversity of the Oral Microbiome in Postmenopausal Women. J Nutr. 2024;154(1):202–212.

18. Fan X, Peters BA, Jacobs EJ, et al. Drinking alcohol is associated with variation in the human oral microbiome in a large study of American adults. Microbiome. 2018;6(1):59.

19. Chaturvedi AK, Vogtmann E, Shi J, et al. Oral Microbiome Profile of the US Population. JAMA Netw Open. 2025;8(5):e258283.

20. Valenzuela MJ, Waterhouse B, Aggarwal VR, Bloor K, Doran T. Effect of sugar-sweetened beverages on oral health: a systematic review and meta-analysis. Eur J Public Health. 2021;31(1):122–129.

21. Li F, Anyaso-Samuel S, Yano Y, et al. Association between diet quality and the oral microbiome in three US cohort studies. J Oral Microbiol. 2026;18(1):2635238.

22. United States Department of Agriculture and United States Department of Health and Human Services. Dietary guidelines for americans, 2020-2025. 2020; 9th:DietaryGuidelines.gov.

23. Subar AF, Thompson FE, Kipnis V, et al. Comparative validation of the Block, Willett, and National Cancer Institute food frequency questionnaires: the Eating at America’s Table Study. Am J Epidemiol. 2001;154(12):1089–1099.

24. Thompson FE, Kipnis V, Midthune D, et al. Performance of a food-frequency questionnaire in the US NIH-AARP (National Institutes of Health-American Association of Retired Persons) Diet and Health Study. Public Health Nutr. 2008;11(2):183–195.

25. Subar AF, Kipnis V, Troiano RP, et al. Using intake biomarkers to evaluate the extent of dietary misreporting in a large sample of adults: the OPEN study. Am J Epidemiol. 2003;158(1):1–13.

26. Subar AF, Midthune D, Kulldorff M, et al. Evaluation of alternative approaches to assign nutrient values to food groups in food frequency questionnaires. Am J Epidemiol. 2000;152(3):279–286.

27. Bolyen E, Rideout JR, Dillon MR, et al. Reproducible, interactive, scalable and extensible microbiome data science using QIIME 2. Nat Biotechnol. 2019;37(8):852– 857.

28. Callahan BJ, McMurdie PJ, Rosen MJ, Han AW, Johnson AJ, Holmes SP. DADA2: High-resolution sample inference from Illumina amplicon data. Nat Methods. 2016;13(7):581–583.

29. Pruesse E, Quast C, Knittel K, et al. SILVA: a comprehensive online resource for quality checked and aligned ribosomal RNA sequence data compatible with ARB. Nucleic Acids Res. 2007;35(21):7188–7196.

30. Faith DP. Conservation evaluation and phylogenetic diversity. Biological Conservation. 1992;61(1):1–10.

31. Shannon CE. A mathematical theory of communication. The Bell System Technical Journal. 1948;27(4):623–656.

32. Lozupone C, Knight R. UniFrac: a new phylogenetic method for comparing microbial communities. Appl Environ Microbiol. 2005;71(12):8228–8235.

33. Li S, Vogtmann E, Graubard BI, Gail MH, Abnet CC, Shi J. fast.adonis: a computationally efficient non-parametric multivariate analysis of microbiome data for large-scale studies. Bioinform Adv. 2022;2(1):vbac044.

34. Barroso E, Martín V, Martínez-Cuesta MC, Peláez C, Requena T. Stability of saliva microbiota during moderate consumption of red wine. Arch Oral Biol. 2015;60(12):1763–1768.

35. Signoretto C, Bianchi F, Burlacchini G, Sivieri F, Spratt D, Canepari P. Drinking habits are associated with changes in the dental plaque microbial community. J Clin Microbiol. 2010;48(2):347–356.

36. Bae J-Y, Seo Y-H, Oh S-W. Antibacterial activities of polyphenols against foodborne pathogens and their application as antibacterial agents. Food Science and Biotechnology. 2022;31(8):985–997.

37. Antezack A, Etchecopar-Etchart D, La Scola B, Monnet-Corti V. New putative periodontopathogens and periodontal health-associated species: A systematic review and meta-analysis. J Periodontal Res. 2023;58(5):893–906.

38. Ikeda E, Shiba T, Ikeda Y, et al. Japanese subgingival microbiota in health vs disease and their roles in predicted functions associated with periodontitis. Odontology. 2020;108(2):280–291.

39. Tagaino R, Washio J, Abiko Y, Tanda N, Sasaki K, Takahashi N. Metabolic property of acetaldehyde production from ethanol and glucose by oral Streptococcus and Neisseria. Scientific Reports. 2019;9(1):10446.

40. Kurkivuori J, Salaspuro V, Kaihovaara P, et al. Acetaldehyde production from ethanol by oral streptococci. Oral Oncology. 2007;43(2):181–186.

41. Sardaro MLS, Grote V, Baik J, Atallah M, Amato KR, Ring M. Effects of Vegetable and Fruit Juicing on Gut and Oral Microbiome Composition. Nutrients. 2025;17(3).

42. Zakis DR, Brandt BW, van der Waal SV, et al. The effect of different sweeteners on the oral microbiome: a randomized clinical exploratory pilot study. J Oral Microbiol. 2024;16(1):2369350.

43. Suez J, Cohen Y, Valdés-Mas R, et al. Personalized microbiome-driven effects of non-nutritive sweeteners on human glucose tolerance. Cell. 2022;185(18):3307–3328.e3319.

44. Gu X, Wang DD, Sampson L, et al. Validity and Reproducibility of a Semiquantitative Food Frequency Questionnaire for Measuring Intakes of Foods and Food Groups. Am J Epidemiol. 2024;193(1):170–179.

