## Supplementary figures and images for "Associations between commonly consumed beverages and the oral microbiome in three US cohort studies"

### Supplemental Figure 1

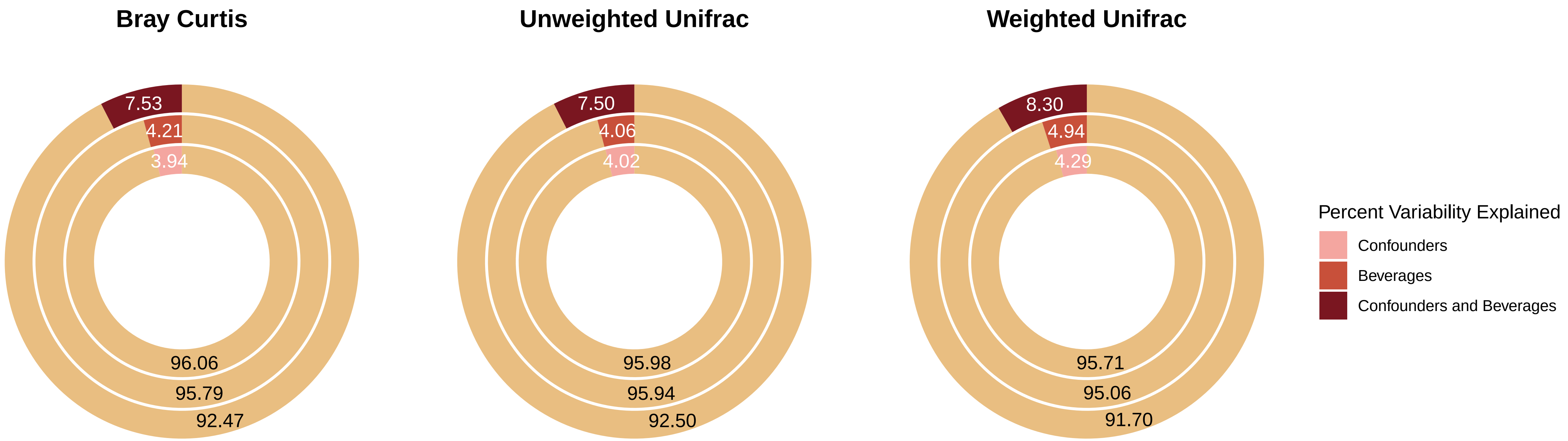
